# Signal detection in the psychotic phenotype: Increased sensory precision and reduced decision threshold associated with psychotic-like experiences

**DOI:** 10.1101/2024.03.14.24304279

**Authors:** Francesco Scaramozzino, Ryan McKay, Nicholas Furl

**Author notes:** Corresponding author: Francesco Scaramozzino; Egham Hill, Egham TW20 0EQ, UK.

## Abstract

**BACKGROUND:** Psychotic-like experiences may reflect disrupted signal detection, whereby individuals detect signals in noisy input that are unlikely to be present. Drawing on predictive coding accounts, we investigated whether increased sensory precision and reduced data-gathering relate to psychotic-like experiences in a signal detection task.

**METHODS:** We fitted drift-diffusion models to Random Dot Motion (RDM) task data completed by 191 participants. Drift rate (proxy of sensory precision) and decision threshold were estimated: 1) across groups differing in psychotic phenotypes, and 2) as outcomes in regression models with psychotic-like experiences as predictors. We also tested whether reduced data-gathering on the beads task replicated prior associations with psychotic phenotypes.

**RESULTS:** Hallucination- and delusion-like experiences were associated with increased drift rates. Hallucination-like experiences also predicted lower decision thresholds. In the beads task, psychotic-like experiences correlated with higher confidence ratings but not with reduced data-gathering.

**CONCLUSIONS:** Our findings indicate that psychotic-like phenomenology is linked to increased precision of signal detection. Overprecise signal detection may increase the likelihood of false positives, potentially leading to anomalous experiences. Alterations of signal detection mechanisms may play a key role in psychosis proneness and, potentially, contribute to the development of psychotic symptoms.

## 1. Introduction

Psychotic phenomenology encompasses a continuum from psychotic-like experiences in health to psychotic disorders^1–4^. Items measuring psychotic phenotype ask individuals about when they inferred information unlikely to be present in their environment or experiences not shared by others (e.g., “Do you ever feel as if people seem to drop hints about you or say things with a double meaning?” ^5^; or “Do you ever detect smells which don’t seem to come from your surroundings?” ^6^). Alterations of signal detection mechanisms may induce the detection of a stimulus (e.g., a sound) when nothing is really there. Signal detection variability in the general population could contribute to having psychotic-like experiences and lead to high scores on psychotic phenotype measurements. Here, we tested whether computational mechanisms of signal detection are associated with several measures of psychotic phenotype in the general population.

Psychotic-like experiences may be linked at the phenomenological, biological and computational levels by predictive coding. Bayesian predictive coding suggests that whether or not we recognise the detection of a signal (e.g., hearing a word in song) depends on the integration of bottom-up and top-down informaiton streams. Within this framework, our experience of reality is constrained by neural inferences that compute signals from the external environment as probability distributions with their own precision (i.e., inverse of variance) ^7–10^. In this view, changes in the encoding of precision by the neural signals could alter perception or beliefs. The predictive coding account of psychosis (PCA)^10^ hypothesises that psychotic experiences may be prompted by less precise priors and/or more precise encoding of sensory information^7,8,10–12^.

This hypothesis has found some support in the context of the beads task (BT). In the BT, participants infer an unknown state of the world (e.g., the dominant colour of beads in a jar) by sequentially sampling pieces of information (e.g., coloured beads). Psychosis has been associated with reduced sampling in the BT, a phenomenon known as jumping-to-conclusions (JTC)^13–17^. In BTs where participants are asked to give their estimates at each piece of collected evidence, new evidence (i.e., a bead of a different colour) impacts probabilistic estimates in schizophrenia patients more than in controls^18–20^. This behavioural pattern is compatible with more precise encoding of sensory information and/or weaker priors^18,19^. At the subclinical level, delusion-like experiences have been associated with a larger weight of sensory evidence in BT^21^ and reduced priors on perceptual inference using the random dot motion task (RDM)^22^.

In this study, we focused on a classic version of the RDM^23^ and investigated the PCA prediction that increased sensory precision contributes to the psychotic phenotype in the general population. Like BT, RDM requires participants to collect evidence from a noisy source to infer a state of the world. We applied drift-diffusion modelling (DDM) to RDM and tested whether psychotic-like experiences were associated with higher sensory precision and less accumulated evidence. The DDM simulates integration of evidence by a decision variable over time^24,25^. In a two-alternative forced choice, the decision variable can reach an upper or a lower threshold (i.e, correct vs. wrong answers). The distance between these threshold values is quantified by the decision threshold parameter *a*, indicating how much evidence is needed to reach a decision. The speed at which the decision variable reaches one of the threshold values is represented by the drift rate parameter, *v*. In DDM, evidence is accumulated by randomly sampling values from the distribution of the information source^24,25^. The precision of the distribution determines the drift rate, that is, the slope of the evidence accumulation process. The more precise the extraction of information, the higher the drift rate.

Notably, the DDM and Bayesian approaches (e.g., PCA) have been considered complementary and formally equivalent ^26–29^. Both DDM and predictive coding involve integrating stochastic information over time to reach a decision (i.e., the threshold values in DDM and the posterior beliefs in Bayesian modelling). Both deal with uncertainty of sensory signals: the DDM does this through varying drift rates, and predictive coding through adjusting the precision assigned to sensory input (see Bitzer et al.^28^ for more details). Bitzer et al. (2014) showed that when Bayesian belief updating is construed as the decision component of a two-alternative forced choice (as we use here), it is equivalent to a DDM. They demonstrated that the drift rate parameter is equivalent to the mean value of the evidence given as input to a Bayesian updating model, approximating the precision of sensory evidence in a predictive coding framework.

Here, we described participants’ performance in terms of the drift rate (*v*) (taken as a proxy for sensory evidence precision) and quantity of accumulated evidence, the decision threshold (*a*). On the basis of PCA and previous findings showing JTC findings in BT^13–17^, we originally expected psychotic phenotypes to correlate negatively with the decision threshold and positively with drift rate in RDM. However, before our data collection, we conducted a pilot study (*N*=20) that showed instead that psychotic phenotypes were associated with lower drift rates and higher decision thresholds, contradicting our initial, more theoretical, expectations. We therefore based our pre-registration on these pilot data (pre-registration link: https://osf.io/d7es8). Nevertheless, we shall show that our main results aligned better with our original theoretical predictions than our pilot data.

## 2 Methods

### Participants

Through Amazon’s Mechanical Turk, we invited a subsample of 200 participants from a larger cohort (N=1002) recruited by Sulik et al.^30^ to take part in our study. We invited only participants who successfully answered at least two of three attention checks. The data from the draws to decision and graded estimates versions of BT (details below) and the Peters et al. Delusion Inventory (PDI)^5^ were collected by Sulik et al. (2023). For the present study, participants performed the RDM and completed the Cardiff Anomalous Perception Scale (CAPS)^6^ and the Aberrant Salience Inventory (ASI)^31^. RDM, CAPS and ASI were submitted through *Gorilla* and undertaken on participants’ computers/laptops. Our study was approved by the ethics committee of Royal Holloway, University of London.

Nine participants did not complete the whole experiment and were excluded. Statistical analyses were hence performed on 191 participants (females=87, males=104; age: M=32.2 yrs, SD=9.8).

### Measures of psychotic phenotypes

We used three measures of subclinical symptoms: the 21-item PDI^5^, measuring delusion-like experiences; the CAPS^6^, measuring hallucination-like experiences; and the ASI^31^, measuring experiences and life attitudes related to aberrant salience (Kapur, 2003). For each measure, we used the total score (sum of subscales). For the between-groups Bayesian hypothesis testing, participants were categorised into two groups for each measure of psychotic phenotypes using the median as a cut-off value; each participant was categorised in three ways: high- or low-PDI group, high- or low-CAPS group and high- or low-ASI group.

### Random Dot Motion Task (RDM)

The visual stimuli for RDM were created through the open-access *JavaScript* code made available on *Codepen.io* by Rajananda et al.^32^. On a black background, we presented 500 white dots, each with a 3-pixel radius. The dots moved at 1 pixel/frame in a circular aperture with a 600-pixel diameter. The dots motion was manipulated by designating “noise” dots moving in randomly assigned directions, and “signal” dots moving in one direction which was either left or right. At each new video frame, the dots were randomly designated to be “signal” or “noise” dots, making it impossible to infer the coherent motion by following a single dot. Each dot was redrawn only after reaching the aperture edges.

We presented in random order: 15 trials of high motion-coherence with 25% signal dots, and 15 trials of low motion-coherence with 15% signal dots. On each trial, the coherent motion was randomly assigned to be left or right. Participants were instructed to use their index fingers and press the “F” button for *left* and the “J” button for *right*. Participants were prompted to answer as quickly and accurately as possible. After each trial, a green tick was displayed for correct answers and a red cross for incorrect. Fast and slow outliers were handled by excluding from analysis trials with reaction times (RT) of <300 msec or >6000 msec^33^.

### “Draws to decision” Beads Task (DTD-BT)

In DTD-BT, participants could draw beads in a predetermined sequence of bead colours (pink and green) in a ratio of 60/40 (sequence: [1- 0- 0- 1- 1- 0- 1- 1- 1- 0- 1- 1- 1- 1- 0- 0- 1- 0- 0- 1]). Participants could decide when to stop sampling and choose the jar from which the sequence was being drawn. If participants wanted to draw more beads than those in the sequence, more draws were randomly generated (up to a maximum of 30). We measured the Draws To Decision scores (DTD), namely the number of beads the participant needed to decide from which jar the beads were drawn. Participants rated their confidence on a 101-point scale ranged from 100% confidence in a “majority pink” urn to 100% confidence in a “majority green” urn at three stages of DTD-BT: at 0 draws (Conf-0), after the first draw (Conf-1) and after the final draw (Conf-N, when they decided to stop data-gathering).

### “Graded-estimate” Beads Task (GE-BT)

In GE-BT, participants were shown a predetermined sequence of 10 beads (ratio: 60/40; sequence: [1, 1, 0, 1, 1, 0, 1, 1, 0, 1]). Using the same scale as for DTD-BT (see section above), participants were asked at each draw how confident they were that the sequence was being drawn from a “majority pink” urn or a “majority green” urn. Here, we computed belief adjustment (ADJ) by measuring participants’ confidence ratings from the disconfirmatory draws. These draws (the 3^rd^, 6^th^ and 9^th^ of each sequence) were disconfirmatory in the sense that they were of a different colour than the majority of preceding draws. From the ratings of these draws, we subtracted the ratings provided in their preceding draws (the 2^nd^, 5^th^ and 8^th^ of each sequence) and then summed these subtractions per sequence to produce the variable ADJ. When the total ADJ score (the overall sum of the subtractions) is positive, it indicates an overall adjustment towards disconfirmatory evidence; if negative, it indicates an adjustment towards the majority colour. We diverged from our pre-registered analysis and took these total ADJ scores as a dependent variable instead of estimates at each draw because we discovered after pre-registration that estimates in response to individual draws were not available from the Sulik et al. dataset.

### Data analysis and computational modelling

Following the data analysis plan we pre-registered, we performed ordinary least squares (OLS) regression models with PDI, CAPS and ASI scores, each the single predictor of one OLS model. For RDM, we also included high versus low coherence as a dummy-coded predictor and used as dependent variables participants’ averages of RT and proportion correct responses. For BT, the dependent variables were DTD, conf-0, conf-1, conf-N and total ADJ scores. Some models showed heteroscedastic residuals (assessed via residuals vs. predicted plots), potentially biasing standard error estimates. To address this and ensure consistency across analyses, we applied quantile (median) regression, which is more robust to OLS assumption violations. Results from these models, reported in the main *Results* section, were consistent with those from the pre-registered OLS models (see *Supplementary Materials*).

These statistical analyses were performed in an *Anaconda Python 3.6* environment using the *statsmodels 12.2* toolbox (https://www.statsmodels.org/dev/index.html). Using the *Tableone* package for Python^34^, we evaluated between-groups differences in distributions of demographic and psychometric variables. We applied the hierarchical DDM (HDDM) to RDM performance using the *HDDM 0.8.0* toolbox^24^ in an *Anaconda Python 3.6* environment. HDDM uses hierarchical Bayesian inference to estimate the posterior distributions of DDM parameters for each participant, allowing them to vary according to condition, group or defined linear regression models. We set the starting point of the sampling (parameter *z*) at 0, simulating unbiased decisions. In all the models we performed, the two choices were coded as correct or incorrect answers, hence *v* can also be considered as an indicator of the RT-accuracy trade-off.

It is uncertain whether a linear increase in psychotic phenotype measures may modulate information processing linearly or only after a threshold score (e.g. median)^35^. We addressed this uncertainty by using two modelling approaches: a between median-split groups approach and a regression model approach, both using Bayesian statistics. For the between-groups approach, we devised HDDMs in which *v* and *a* parameters could vary between conditions and groups (see *Table 1*). Here, we report these HDDMs instead of those proposed in the pre-registration. The pre-registered models evaluate only one motion-coherence condition at the time not accounting for within-participant variability between conditions and compromising the precision of parameter estimations. These pre-registered models showed results consistent to those from the models presented here (see *Supplementary Materials*).

**Table 1.** HDDM specifications. Conditions/groups according to which drift rate and decision threshold parameters were free to vary. HC=High coherence LC=Low coherence LR=Linear Regression

| Between-group Models | Condition/ Groups | Regression Models | Predictors |
| --- | --- | --- | --- |
| Null-model | None | Null-model | None |
| HDDM-C | HC vs. LC | HDDM-LR-C | HC vs. LC |
| HDDM-C-PDI | HC vs. LC \|<br>Low-PDI vs.<br>High-PDI | HDDM-LR-C-PDI | PDI \| HC vs. LC |
| HDDM-C-CAPS | HC vs. LC \|<br>Low-CAPS vs.<br>High-CAPS | HDDM-LR-C-CAPS | CAPS \| HC vs. LC |
| HDDM-C-ASI | HC vs. LC \|<br>Low-ASI vs.<br>High-ASI | HDDM-LR-C-ASI | ASU \| HC vs. LC |

For the regression model approach, we used the *HDDMregressor* class of the HDDM package to run DDMs in which *v* and *a* parameters varied according to a Bayesian linear regression model specified in *Table 1.* We z-scored the values of all continuous predictors before running the models with *HDDMregressor*.

For the between-group models, we compared posterior probability distributions of parameters between phenotype groups within each motion-coherence condition. We computed the probability that a parameter is higher or lower in one condition/group than in the other. For the Bayesian regressions, we evaluated whether the probability *P* of the posterior probability distribution of the regression coefficient *ß* was significantly different from 0, meaning that at least 95% of the distribution was either >0 (positive *ß*) or <0 (negative *ß*)^24^. To maintain a notation familiar to the frequentist approach, we considered our results significant when the probability of the disconfirming hypothesis was <0.05 ^24^.

We evaluated model fit using the Deviance Information Criterion (DIC). DIC penalises models for complexity and lower values indicate a better fit. We compared each model to a null model and to models accounting only for motion-coherence in RDM (HDDM-C, HDDM-LR-C), considering a difference (Δ) of 10 DIC points as significant^36^. Model reliability was further tested through data simulation and parameter recovery (see *Supplementary Materials*). For each DDM, Markov chain Monte Carlo simulations were used to generate 20,000 samples. The first 2,000 samples were discarded as burn-in, a thinning factor of 5 was used and outliers were set at 10%. The convergence of Markov chains was assessed by visual inspection and by computing the Gelman-Rubin statistic and verifying values ranging between 0.9 and 1.1.

## 3 Results

### HDDM modelling: Effects on drift rate and decision threshold

#### Convergence

All models showed evidence of convergence from visual inspection and the Gelman-Rubin diagnostic, the values of which ranged between 0.9 and 1.

#### HDDM between-group analysis

*Table 2* shows DICs for between-group models. The *HDDM-CAPS-C* is the only model showing significantly better DIC when compared with both the *Null-model* and the *HDDM-C*, indicating that incorporating both motion coherence and the median-split grouping of hallucinatory phenotype (low- and high-CAPS) significantly improves the model.

**Table 2.** DIC between-group HDDMs. *The second column shows values of Deviance Information Criterion (DIC) for each model in descending order (lower DIC, better fit); The third column shows differences in DIC (*Δ *DIC) between each model and the* Null-model (parameters not varying for any variable)*; The last column shows* Δ *DIC between each model and the* C-model (parameters varying for motion-coherence only).

| | DIC | $\Delta$ DIC Null-model | $\Delta$ DIC HDDM-C |
| --- | --- | --- | --- |
| HDDM-CAPS-C | 16067.09 | -96.1* | -13.47* |
| HDDM-ASI-C | 16078.93 | -84.26* | -1.63 |
| HDDM-C | 16080.56 | -82.63* | 0.0 |
| Null-model | 16163.19 | 0.0 | 82.63 |
| HDDM-PDI-C | 16188.75 | 25.56 | 108.19 |

\*DIC significantly lower ( $\Delta$ DIC < -10)

**Table 3.** DIC regression HDDMs. *Values of Deviance Information Criterion (DIC) for each model in descending order (lower DIC, better fit); differences in DIC (*Δ *DIC) between each model and the* Null-model *(parameters not varying for any variable);* Δ *DIC between each model and the* C-model (parameters varying for motion-coherence only).

| | DIC | $\Delta$ DIC Null-model | $\Delta$ DIC HDDM-C |
| --- | --- | --- | --- |
| HDDM-LR-CAPS-C | 16050.72 | -112.47* | -29.75* |
| HDDM-LR-PDI-C | 16062.89 | -100.3* | -17.58* |
| HDDM-LR-C | 16080.47 | -82.72* | 0.0 |
| HDDM-LR-ASI-C | 16083.34 | -79.85* | 2.87 |
| Null-model | 16163.19 | 0.0 | 82.72 |

\*DIC significantly lower ( $\Delta$ DIC < -10)

Although *HDDM-ASI-C* outperformed the *Null-model,* it did not show a better DIC than the *HDDM-C*, suggesting that ASI grouping does not contribute to explaining the data further than taking only motion coherence into account. In the following, we report the between-group differences in posterior probability estimates from *HDDM-CAPS-C,* the model for which phenotype successfully explained participants’ behaviour.

*Figure 1* shows results from *HDDM-CAPS-C*. For each group, drift rates were significantly higher in high motion-coherence than in low motion-coherence (see *Fig. 1-A*), reflecting the noise in the physical stimulus (percentage of coherently moving dots), thus empirically confirming that drift rate can be considered a good proxy for precision of sensory evidence^25^. In parallel, decision threshold was also significantly higher in high motion-coherence than in low motion-coherence (see *Fig. 1-B*).

**Fig. 1.**
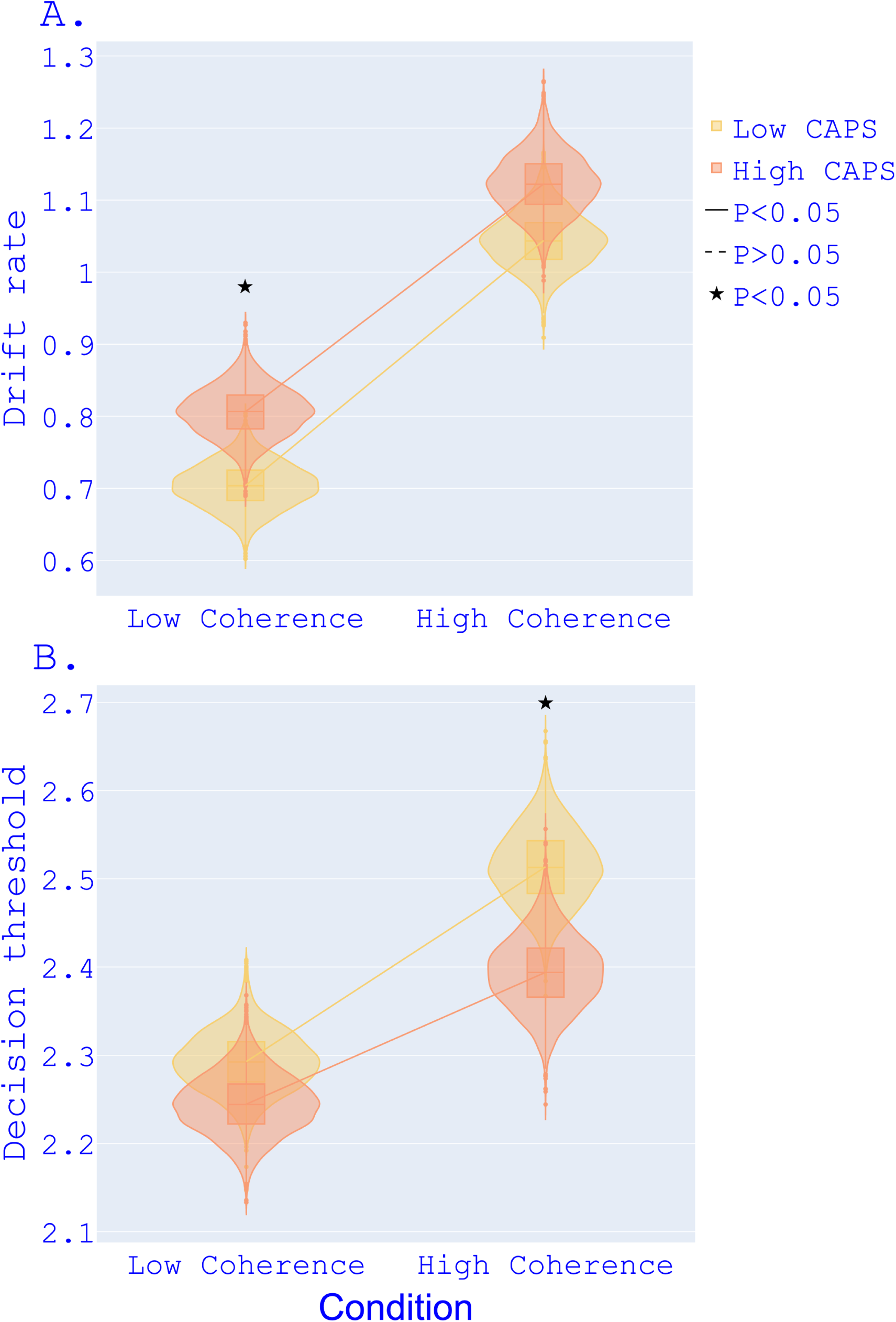
Between-group comparison. Posterior probability density distributions of drift rate (A) and decision threshold (B) for median-split groups of psychotic phenotypes. The ends of the boxes show the lower and upper quartiles of the distributions, while the line inside the box marks the median.

In low motion-coherence, the high-CAPS group showed significantly higher drift rates than the low-CAPS group (see *Fig. 1-A, left*). An analogous result was present as a trend in high motion-coherence (see *Fig. 1-A, right*). The high-CAPS group also showed a significantly lower decision threshold than the low-CAPS group in high motion-coherence (see *Fig. 1-B*). Parameter recovery reproduced these results showing reliability for these model estimations (see *Supplementary Materials*).

### HDDM regression analysis

In regression HDDM, DDM parameters were set to vary as response variables of linear regressions. The models where motion-coherence and either PDI or CAPS were predictors (*HDDM-LR-PDI-C*, *HDDM-LR-CAPS-C*) showed significantly better (i.e., lower) DICs compared to the *Null-model* and *HDDM-LR-C,* which included coherence as the only predictor. In contrast, *HDDM-LR-ASI-C* did not outperform *HDDM-LR-C*. The regressions confirmed the results from the between-group analysis and showed CAPS predicting higher drift rates and lower decision thresholds (see *Figure 2*, left). For PDI, the regressions showed a positive effect of PDI on drift rate that did not emerge from the between-group analysis (see *Figure 2*, upper right). Parameter recovery reproduced these results (see *Supplementary Materials*).

**Fig. 2.**
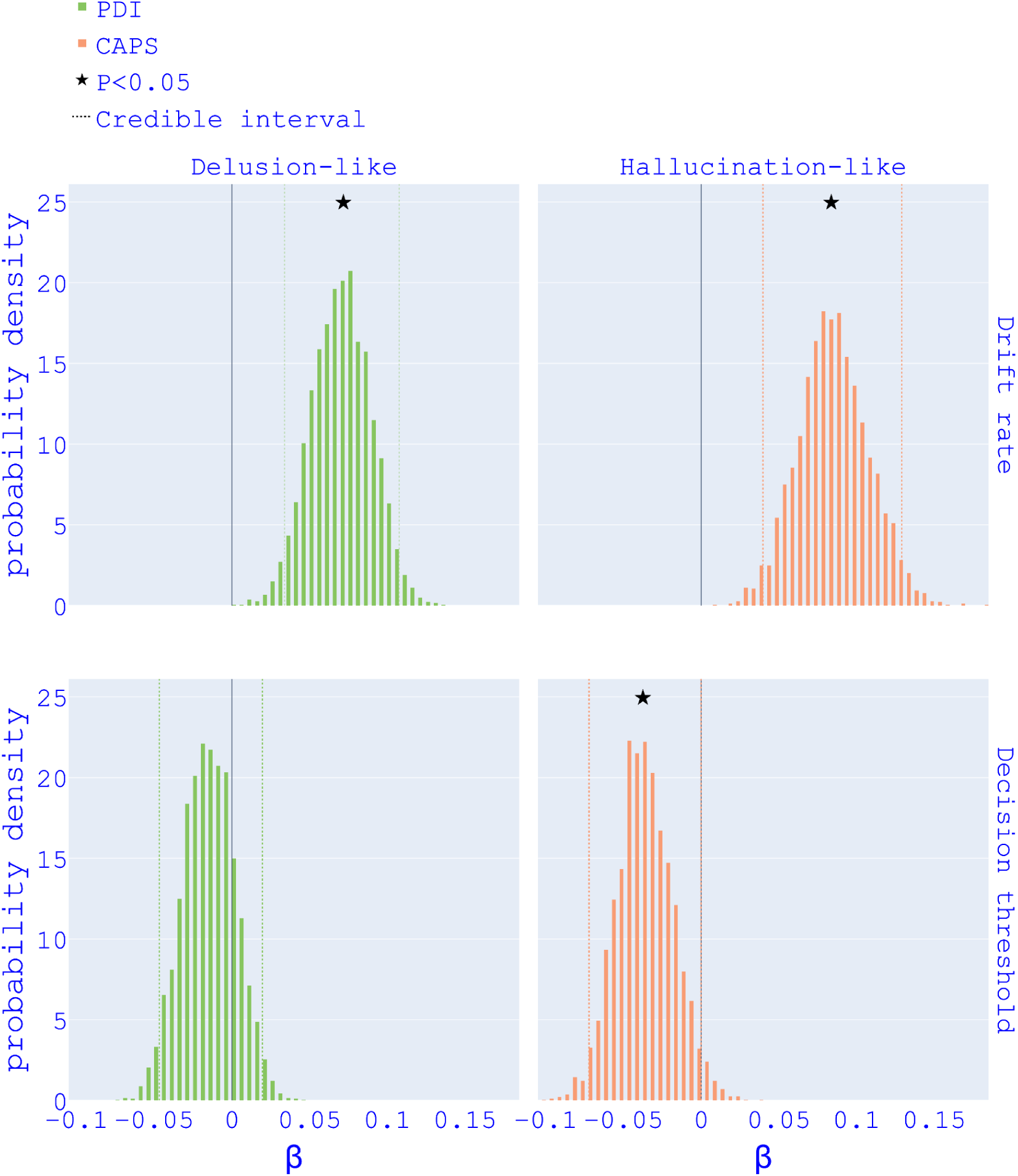
HDDM regressions. Posterior probability density distributions of *ß* coefficients for PDI and CAPS respectively from HDDM-LR 4 and HDDM-LR 5.

### RDM RTs and accuracy

The quantile regressions (*Figure 3*) did not show any effects of psychotic phenotype measurements on the average values of RT or response accuracy.

**Fig. 3.**
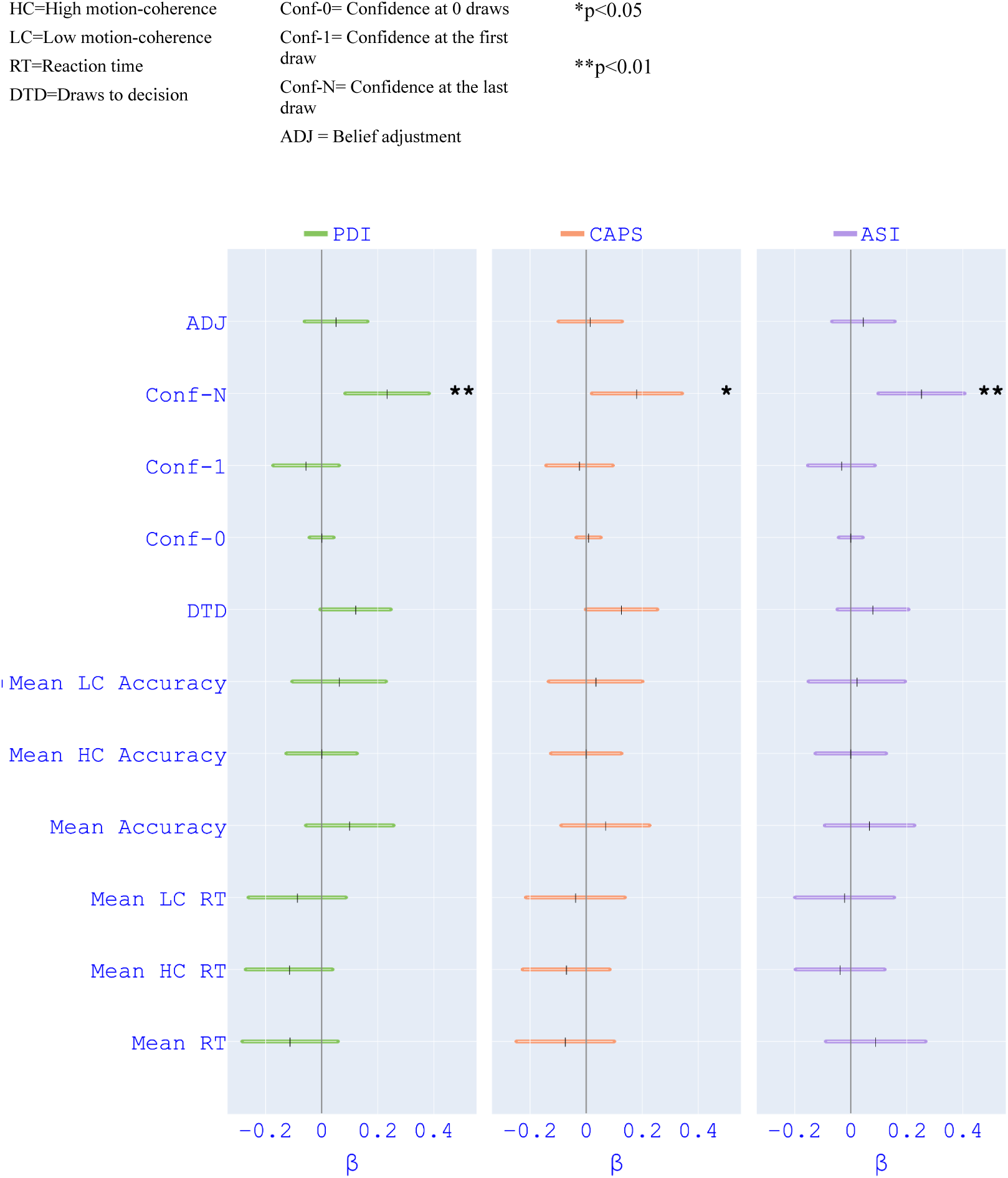
Quantile regression models. Results from quantile (median) regression models with PDI, CAPS or ASI as the predictor. “Mean Accuracy” indicates the percentage of correct responses. Vertical dashes indicate the estimates for *β*. Coloured points indicate the upper and lower bounds of confidence intervals.

### Probabilistic reasoning: BT

Although we found no relationship between psychotic phenotypes and DTD or ADJ (see *Fig. 3*), the results from the quantile regressions showed a positive effect of PDI, CAPS and ASI on Conf-N (i.e., confidence rating at the final draw, see *Fig. 3)*.

### Demographics and Subclinical psychosis

In *Table 4*, we report the main demographics of our participant sample and the means and quartiles of the distributions for high and low PDI, CAPS, and ASI groups. Only between low-CAPS and high-CAPS groups was there a significant difference in age with participants in the high-CAPS group being older (*t* (189) = −0.36, *p*<0.05). In all group pairs, we see an uneven distribution of the psychotic phenotype measures: CAPS and ASI scores are higher for the high-PDI group, PDI and ASI scores are higher for the high-CAPS group, PDI and CAPS scores are higher for the high-ASI group.

**Table 4.**
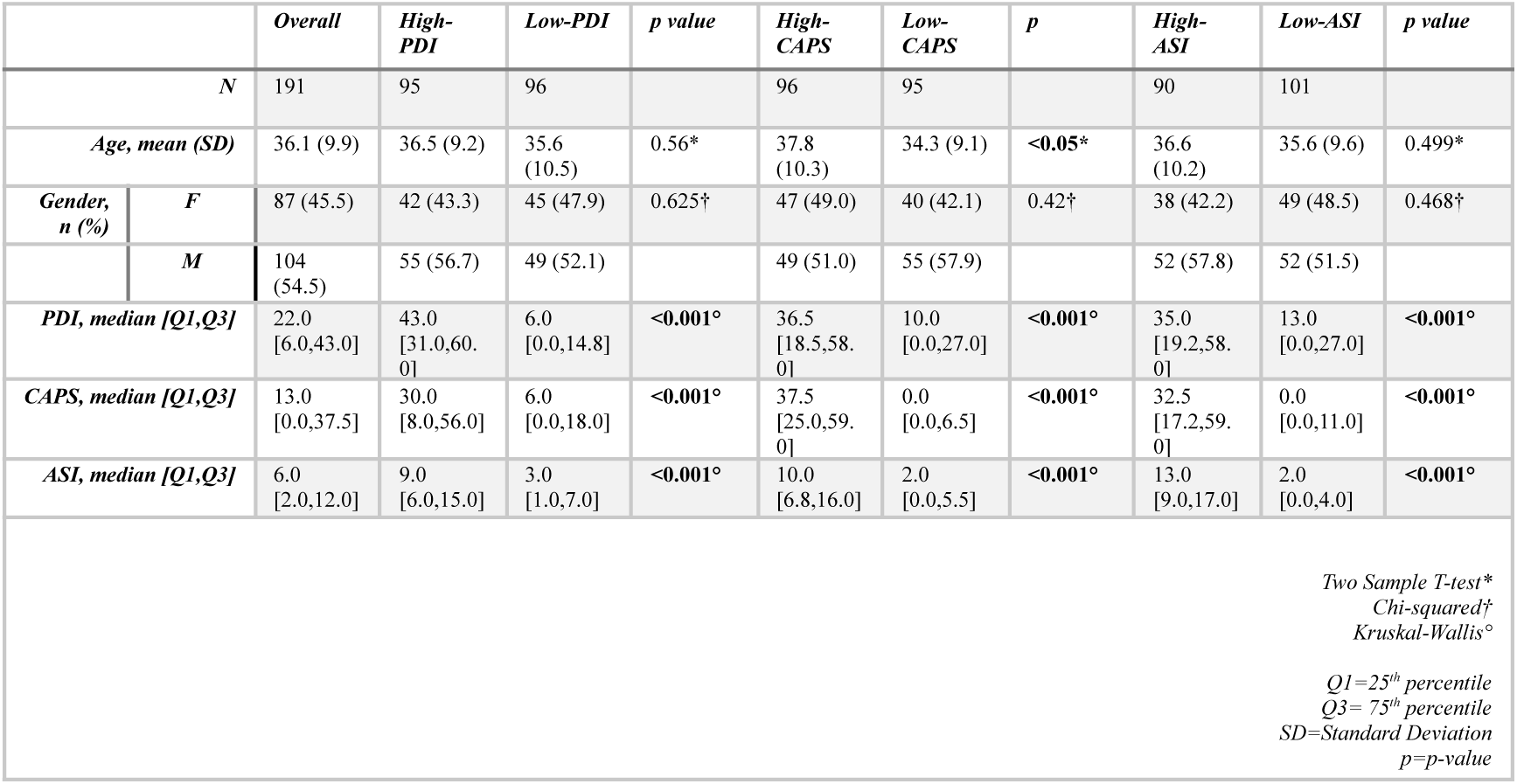
Differences between psychotic phenotype groups. Median values of demographic, psychometric and psychotic trait variables with tests of their group differences (significant differences in bold).

## 4 Discussion

We used a classic RDM task ^23^ and DDM to test whether psychotic-like experiences were linked to increased drift rate (proxy for sensory precision) and reduced decision threshold. Our group analysis results showed that hallucination-like experiences (i.e., CAPS) were associated with both increased drift rate (at least for challenging conditions) and decreased decision threshold. The regression modelling confirmed this effect and showed delusion-like experiences associated with higher drift rates, backed up by a reliable parameter recovery and accurate prediction of participants’ RTs (see *Supplementary Materials*).

### Psychotic-like Experiences and Perceptual Inference

Overall, our findings suggest that increased precision in signal detection is present in members of the general population with higher psychotic phenotypes. This variability in information processing may be linked to a predisposition to developing a psychotic disorder or simply contribute to the emergence of psychotic-like experiences without any clinical implications. Future studies comparing psychotic phenotype in the general, at-risk and early-onset psychosis populations may further investigate the link between the DDM patterns we found and the evolution of pathological symptoms.

Median split group analyses and regression analyses largely agreed on effects of CAPS, though regression showed additional effects of PDI. One potential explanation for the discrepancy for delusion-like experiences is the difference in statistical power between the two methods. The between-group comparison based on the median split may have lacked sufficient power to detect the effects of PDI on drift rate. Additionally, the median split may not represent a critical threshold for the impact of PDI on task performance. The effect of PDI on drift rate may operate along a continuum, where individual differences across the entire range of PDI scores are informative.

Previous studies have associated psychotic-like experiences with increased influence of prior beliefs^37–40^. These studies involved the discrimination of visual stimuli, including pictures of humans and other animals in static noisy pictures^37–39^. Our findings do not contrast with this literature but suggest that when discriminating motion of socially neutral stimuli, the psychotic phenotype is characterised by increased precision of sensory evidence. Our paradigm did not include any manipulation of prior beliefs on the direction of coherent moving dots, hence we cannot make any inference on participants’ encoding of priors. Studies inducing prior beliefs in RDM found PDI associated with weaker perceptual priors^22^ and patients with schizophrenia to be less prone to perceive prior-induced dot motion^41^. Together these findings as well as ours suggest that the balance of prior versus sensory precision varies across psychotic phenotypes depending on whether pictorial stimuli are socially meaningful or socially neutral, as well as static or in-motion. These differences may be indicative of differential top-down/bottom-up precision modulations throughout the perceptual processing hierarchy. Such differences are in line with the PCA hypothesis that sees N-methyl-D-aspartate receptor hypofunction as a driving pathogenic mechanism disrupting higher-levels priors and enhancing bottom-up sensory signalling^10^. Future studies might focus on the link between N-methyl-D-aspartate receptor hypofunction and differential precision encoding according to stimulus’ nature (socially meaningful vs. neutral; static vs. in-motion).

We found that individuals with higher CAPS scores required less evidence (i.e., had lower decision thresholds) to commit to a decision. Notably, requiring less evidence did not impair accuracy. Participants with a higher hallucinatory phenotype not only required less information but also gained sensory signals more quickly (higher drift rates) compared to those with a lower hallucinatory phenotype. These results may suggest an advantage in signal detection associated with the hallucinatory phenotype, at least in the context of the RDM. The same mechanisms enhancing signal detection could also contribute to hallucination-like experiences. We can speculate that when confronted with noise in the absence of a signal, this enhanced signal detection capacity might also lead to the detection of false positives. This hypothesis is consistent with evidence showing auditory hallucinations in healthy voice-hearers to be linked to higher rates of false positives in auditory signal detection tasks^42,43^. Similarly, hallucination-like experiences have been associated with higher rates of false positives in word recognition tasks^44^. A version of RDM designed to measure false positive rates might test this hypothesis in the future.

In contrast to PDI and CAPS, aberrant salience as measured by ASI did not show any effect on DDM parameters. Like PDI and CAPS, ASI is also considered to measure a psychotic trait^31^ and in our study, all three measures were positively associated with each other (see *Table 2*). This discrepancy in the impact on RDM performance between ASI and the other subclinical traits might represent a dissociation between core psychotic-like symptoms (delusional ideation and anomalous perceptions) and aberrant salience of stimuli.

### Psychotic-like Experiences and Cognitive Inference

We found all three measures of psychotic phenotype to be associated with higher confidence ratings at the final urn choice in DTD-BT. These results are compatible with findings indicating general metacognitive deficits in both patients with schizophrenia^15^ and subclinical psychosis^45^. In contrast, we did not find statistical evidence for any relationship between psychosis phenotype and DTD, which is inconsistent with findings showing associations between psychotic phenotype and JTC. Different studies have pointed out how other factors modulate sampling in BT and possibly create an illusory positive association between JTC and delusions: miscomprehension of the task^46^, motivation^47^, socio-economic status^48^ or general cognitive ability^49^. Ross et al. (2016)^50^ found that PDI had no effect on DTD, and showed that cognitive style (analytical vs. intuitive) could be considered a more reliable predictor of sampling behaviour in BT. These results illustrate the sporadic nature of the association between delusion-like beliefs and reduced sampling in the BT^14^ and show how alterations of perceptual inference (e.g., on the RDM task) might represent instead a more prominent feature of subclinical psychosis.

We also did not find any significant relationship between psychotic-like experiences and ADJ. Although this BT behavioural pattern has been shown in schizophrenia patients with delusions^18–20^, it has repeatedly failed to be replicated in individuals from the general population with high scores for delusional ideation^30,46,51^. Our study therefore supports the idea that ADJ does not vary with measures of subclinical psychosis.

### Limitations

A few points need to be considered when interpreting our results. First, the HDDM regression effect sizes we observed are relatively small but consistent with the effect sizes found in the literature on individual differences in the psychotic phenotype^14^. Second, as mentioned earlier, we did not measure prior beliefs. Future paradigms could address this by introducing dot direction biases to assess the influence of prior encoding on perceptual inference and better characterise participants’ performance. A limitation of our and many Beads Task (BT) paradigms is their one-trial design, given that JTC effects diminish over repeated trials.^52^. It remains possible that a repeated-measures BT could have revealed a negative association between DTD and psychotic-like experiences. Lastly, it is important to note that PDI and BT data were collected at different times from CAPS, ASI, and RDM data. While we found consistency in measures of psychotic phenotype across median-split groups, we cannot entirely rule out the possibility that our results might be different if we collected all measures at the same time.

In summary, our study highlights an association of both hallucination- and delusion-like experiences with increased sensory precision in detecting motion of socially neutral stimuli. A higher hallucinatory phenotype was also associated with a decreased decision threshold. We did not find any significant behavioural pattern associated with psychotic-like experiences in cognitive inference (i.e., on the draws to decision beads task). More broadly, our findings suggest that alterations in precision encoding of sensory information might be implicated in psychotic phenomenology in the general population, in line with the predictions of PCA. Investigating whether and how the neurocomputational specificities we found at the subclinical level vary in the clinical population could shed light on mechanisms driving the formation of full-blown psychotic symptoms.

## Acknowledgements

This work was supported by a grant to R.M. from the NOMIS Foundation (“Collective Delusions: Social Identity and Scientific Misbeliefs”).

## Data Availability Statement

The data that support the findings of this study are openly available at: https://github.com/frscaramozzino/Signal_detection_within_psychotic_phenotypes.

## Supplementary materials

### 1. Distributions of psychotic phenotype measures

For a full display of the psychotic phenotype data in our sample, we report here the medians (red vertical lines) and the frequency distributions in sample percentage for PDI, CAPS and ASI (*Figure S1-A-B-C*).

**Fig S1.**
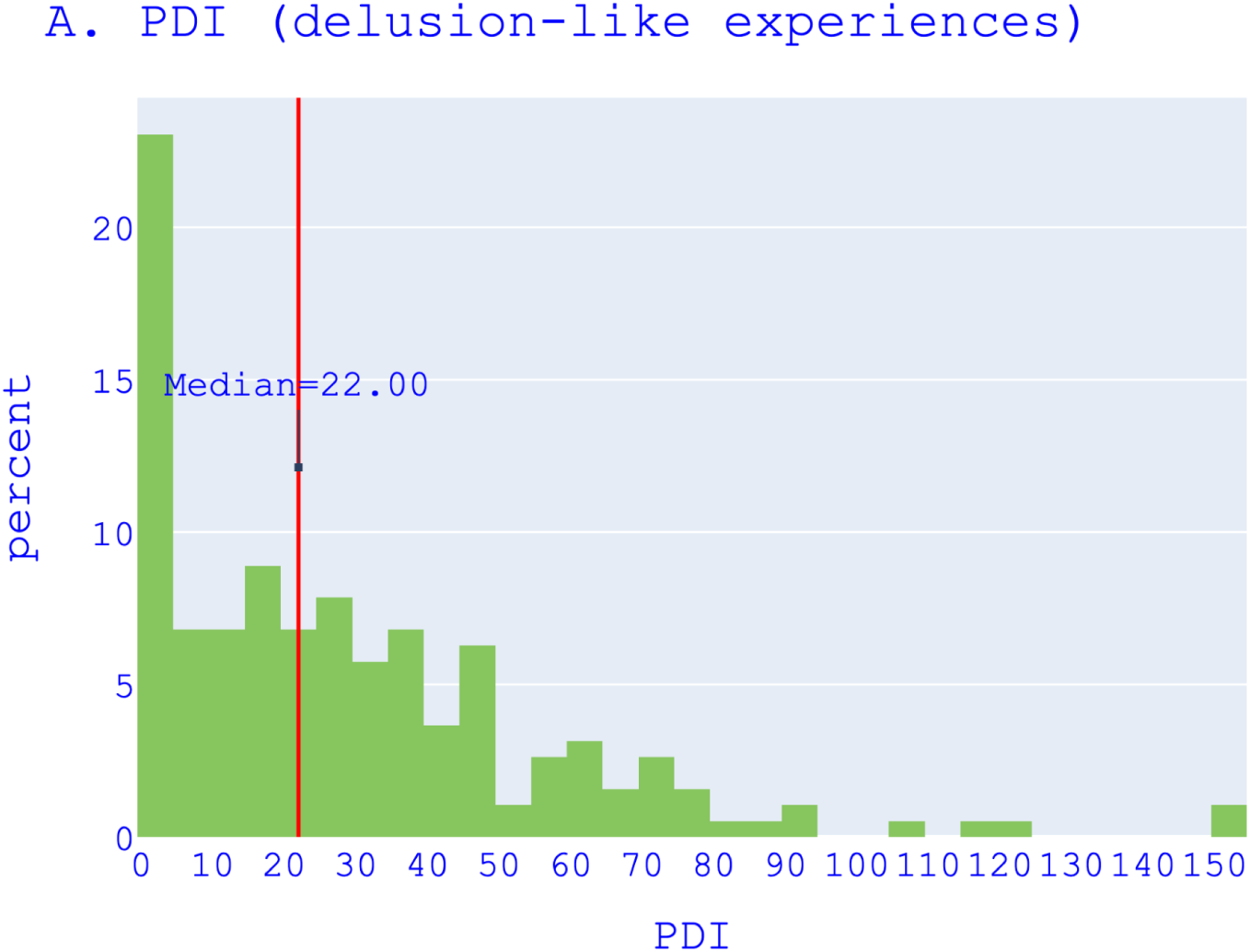
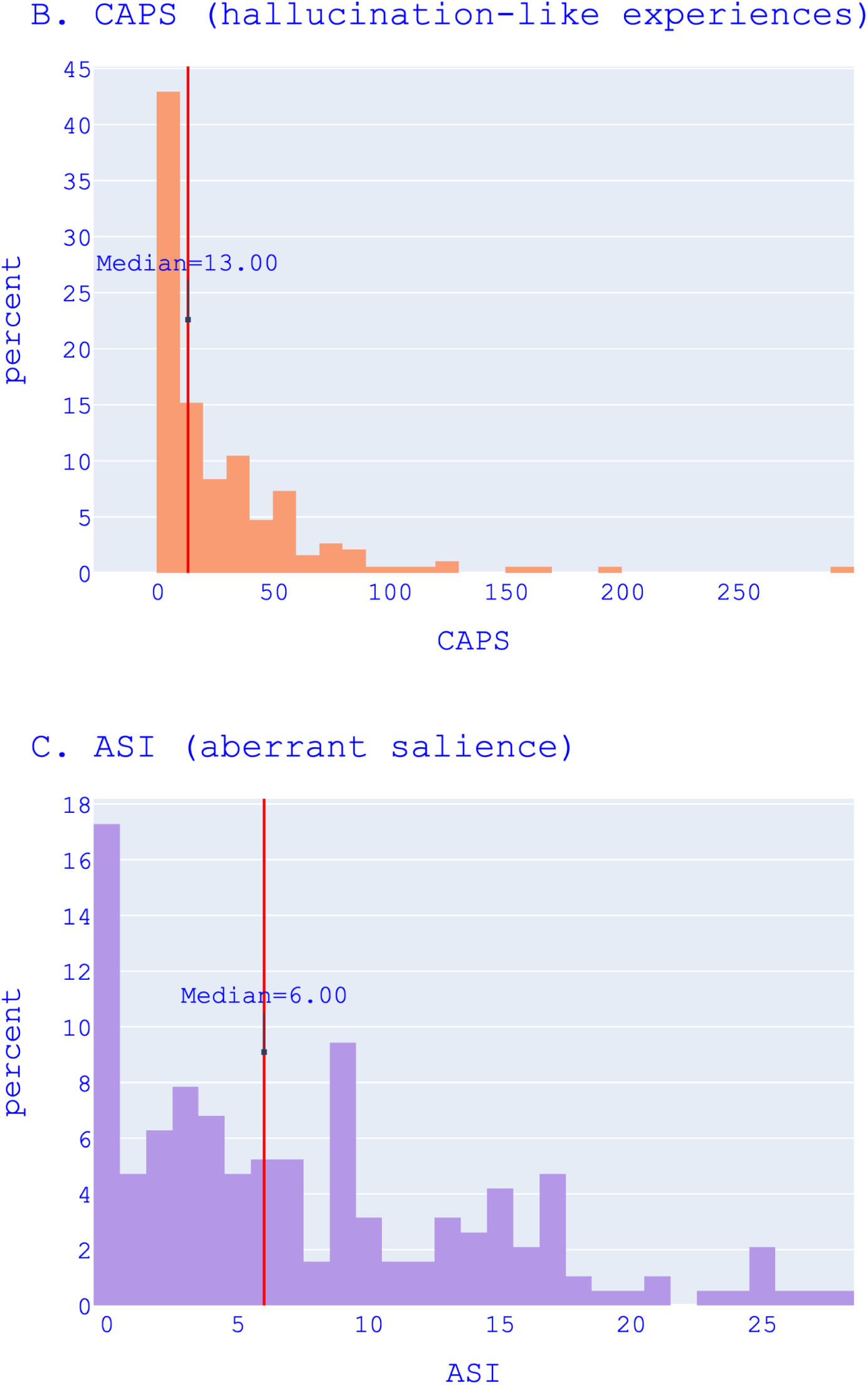
Frequency distribution of psychotic phenotype measures. Distribution in participant percentage of (A) Peters et al. Delusion Inventory (PDI), (B) Cardiff Anomalous Perceptions Scale (CAPS) and (C) Aberrant Salience Inventory (ASI).

### 2. Data Simulation and Parameter Recovery

We provide here a comprehensive overview of the performance of the hierarchical drift-diffusion models (HDDMs) best-fitted to the participant choices from the Random Dot Motion task (RDM). Following good practice guidance in computational modelling of behaviour (Vandekerckhove & Tuerlinckx, 2007; Wiecki et al., 2013; Wilson & Collins, 2019), we assessed the validity of our models through two main steps: simulation of data and recovery of parameters. By evaluating the performance of our models, we also evaluated the relative merits of different modelling techniques, including median split versus regression techniques with a view to developing modelling guidance for future studies. Here, we present a qualitative comparison between observed and simulated data, followed by the recovery of the estimated parameters. In *Section 1*, we present the methodology we used, and in *Sections 2* and *3* the results of data simulation and parameter recovery. Finally, in the *Discussion* Section, we comment and draw conclusions from *Sections 2* and *3*.

#### 2.1 Methodology

##### Data simulation

We used the *hddm.generate.gen_rand_data(…)* function of the *HDDM 0.8.0* package (Wiecki et al., 2013) to simulate reaction times (RTs) and correct and error responses given the mean and standard deviation of DDM parameters. The function allows specification of the values of parameters for different discrete levels (e.g., conditions or groups), and reproduces the size and structure of the data in terms of the number of trials and participants per level. For between-group HDDMs, we inputted the mean and standard deviation of the fitted parameters for each condition/group level and obtained RTs and responses for each group/condition.

For regression HDDMs, more steps than for the between-group HDDMs were needed to simulate data sets that reproduce the structure of the observed ones. In fact, in the regression HDDMs, we had a two-level dummy coded variable (motion-coherence) and a continuous variable (PDI or CAPS) predicting each parameter of interest (drift rate or decision threshold). One obstacle in the data simulation was to inform the simulation function of the effect of the continuous variable on the parameters. In fact, *hddm.generate.gen_rand_data(…)* simulates data given discrete parameter values. We approached this problem by computing a mean predicted value of parameters for each level of the dummy coded variable including the regression coefficients of the continuous variable in each level as shown below in *Equation 1* and *Equation 2*.

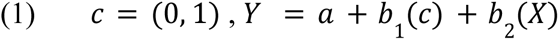

In *Equation 1*, *Y* denotes the vector of predicted values (e.g., drift rate) for the level of the dummy coded variable *c* (*c=0* ∨ *c=1*), *X* the continuous variable predictor (e.g., PDI or CAPS) and *a* the intercept. We computed *Y* for both *c=0* and *c=1* and obtained two vectors, one for each level of the dummy coded variable, of the predicted parameters *Y_0_* and *Y_1_*. We then averaged the values for each array and obtained the average *Y_0_* for level 0 and average *Y_1_* for level 1, which are the mean predicted parameters to be inputted to *hddm.generate.gen_rand_data(…)*. To simulate the models, we needed to include in the simulated dataset a value of the continuous variable (PDI and CAPS) per simulated participant. Since the data were simulated employing a stochastic approximation of the effect of the continuous variable on the parameters (i.e., the mean predicted value), we took a similarly stochastic approach and pooled randomly from our empirical data values of PDI and CAPS and assigned them to the simulated participants.

To capture how well the models whose results we reported predict participants’ raw data, we simulated 100 datasets and averaged the 10th, 30th, 50th, 70th and 90th quantiles of the simulated RT distributions for correct and error responses. We then plotted in a linearity plot (or QQ plot, shown in *Figure S3* and *Figure S5*) the observed RT quantiles on the x-axis and the simulated quantiles on the y-axis to obtain a line that we contrasted with an “optimal simulation line” drawn by plotting observed quantiles on both axes. To quantify the distance between the simulated and the observed quantiles, we took as metrics the root-mean-squared error (RMSE) and the Standardised Mean Difference (SMD; also known as Cohen’s D). RMSE is a measure of the average difference between predicted and observed values and it is expressed in the units of the variable value. The SMD is the ratio of the difference between the means and the sum of the standard deviations of the two distributions. Unlike the RMSE, the SMD is a standardised metric suitable for comparing the prediction performance across different magnitudes of RTs (e.g. between correct and error responses, the latter usually slower).

##### Parameter recovery

We performed the parameter recovery for between-group and regression HDDMs reported in the *Results* section. In the between-group HDDMs, drift rate and decision threshold were allowed to vary for motion-coherence (high vs. low) and median-split groups. In the regression models, drift rate and decision threshold were free to vary as the response variable of a linear regression model with motion-coherence and PDI (HDDM-LR-PDI-C), CAPS (HDDM-LR-CAPS-C) or ASI (HDDM-LR-ASI-C) scores as predictors. We refer here to these models as *empirical models*, because their parameters were estimated from fits to human RTs and accuracy rather than to simulated data. For each empirical model, we simulated one dataset (following the procedures described in the preceding paragraph) and fitted an equivalent *simulation model*. We evaluated the distance between the distributions of the simulated and empirical parameters using RMSE and SMD. For between-group HDDMs, we computed RMSE and SMD between the estimated and simulated distributions of DDM parameters per group (e.g. low-PDI) or condition (e.g. high motion-coherence). For regression HDDMs, we computed the RMSE and SMD between the empirical and simulated *β* coefficients. Lower RMSE and SMD between estimated and simulated parameters indicate a better recovery and a more precise estimation of the true parameters for groups, conditions or predicted variables. Additionally, we considered whether the simulation models could reproduce the DDM parameter differences and *β* estimates reported in the results of the empirical models. If the simulated parameters adequately recover the effects observed, it suggests that the model is accurately capturing the processes that produced the empirical data.

Overall, the recovery of parameters is an essential step in validating computational models of behaviour as it helps to establish the reliability, validity, and generalizability of the model’s results, and increases confidence in the model’s ability to accurately describe and explain data from different samples of the same population (Vandekerckhove & Tuerlinckx, 2007; Wiecki et al., 2013; Wilson & Collins, 2019).

#### 2.2 Data simulation and parameter recovery results

Here, we report the data simulation and parameter recovery for between-group and regression HDDMs. In *Figure S2*, we show the distribution of RTs inputted to these models.

**Fig. S2.**
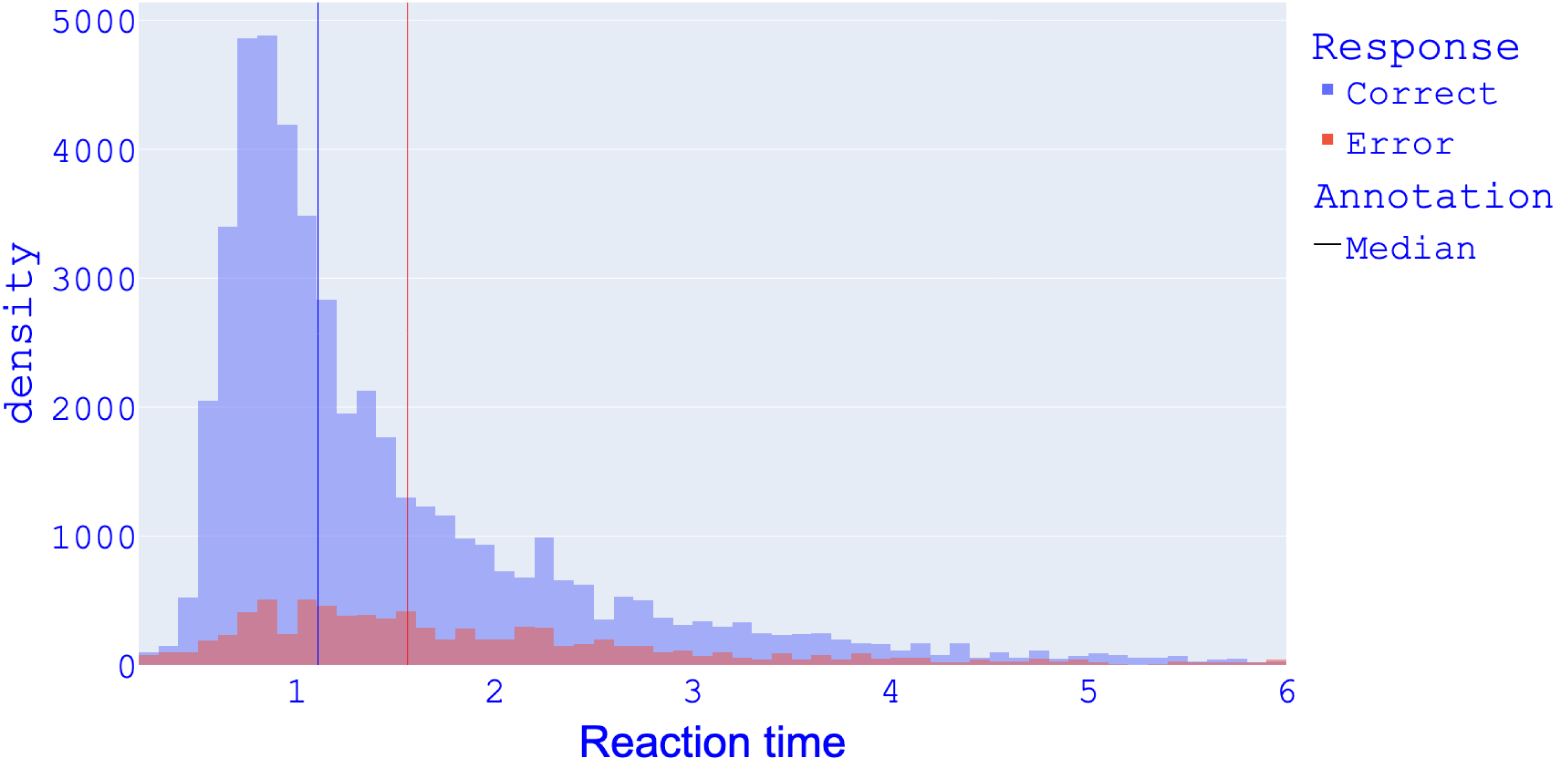
RT distribution Study1. Distribution of reaction times (in seconds) for correct and error responses as inputted to HDDMs in *Study* 1; fast outliers cut-off: 0.2 s; slow outliers cut-off: 6 s.

##### Between-group HDDMs

*Figure S3* plots the quantiles of simulated RT distributions against the observed RT quantiles for the between-group winning model HDDM-CAPS-C.

**Fig. S3.**
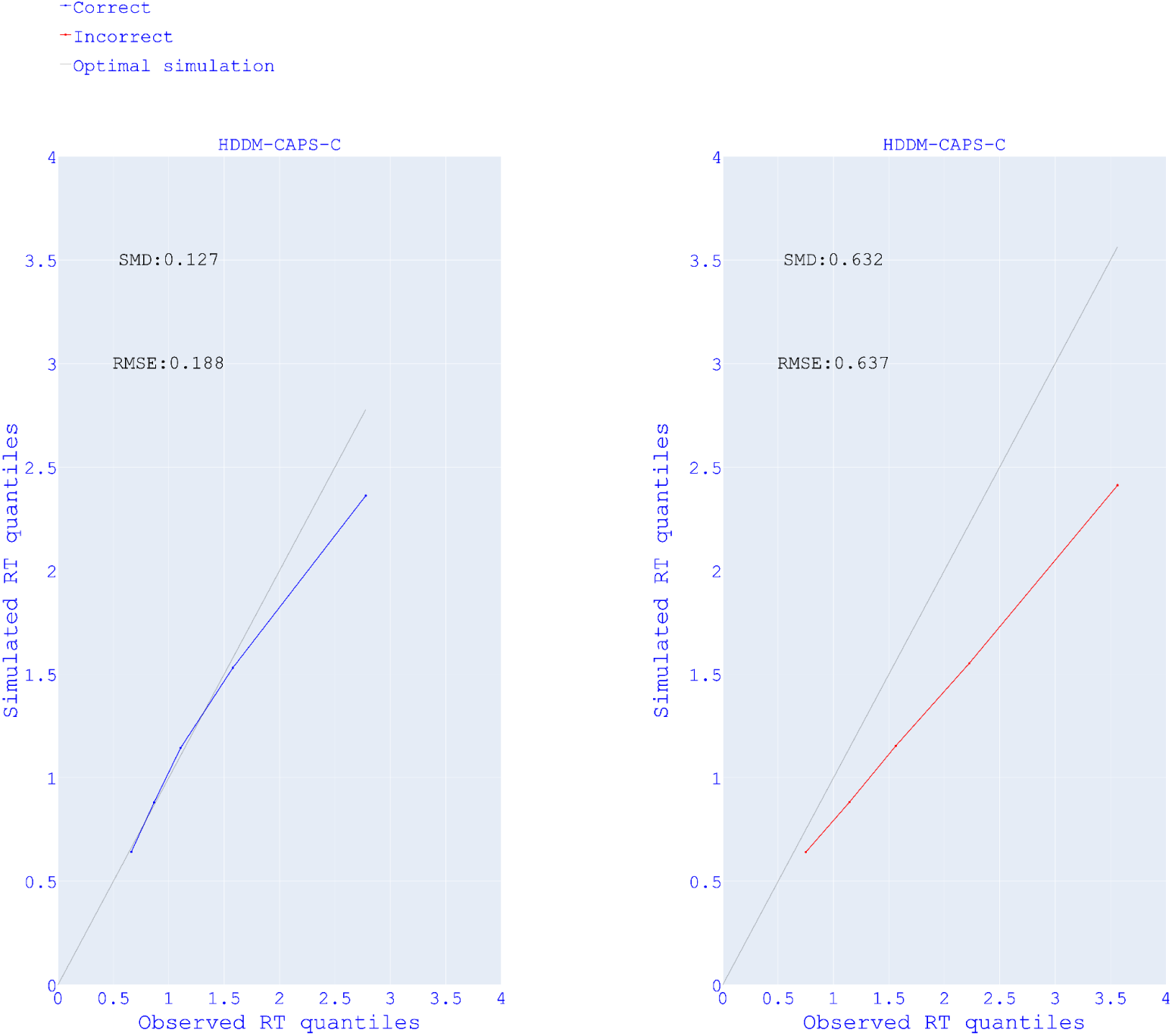
RT predictions of between-group HDDMs Study 1. Linearity plots of observed reaction time (RT) quantiles (x-axis) and simulated RT quantiles (y-axis) for correct and error responses of between-group models in *Study 1*. SMD= Standardised mean difference; RMSE= Root mean squared error.

In *Figure S4*, note that simulated parameters tend to be higher than the estimated parameters. This overestimation did not prevent the recovery of the significant difference in parameter estimates between CAPS median-split groups shown by the empirical model (see: *Figure S4* top-left and *Figure S4* bottom-right).

**Fig. S4.**
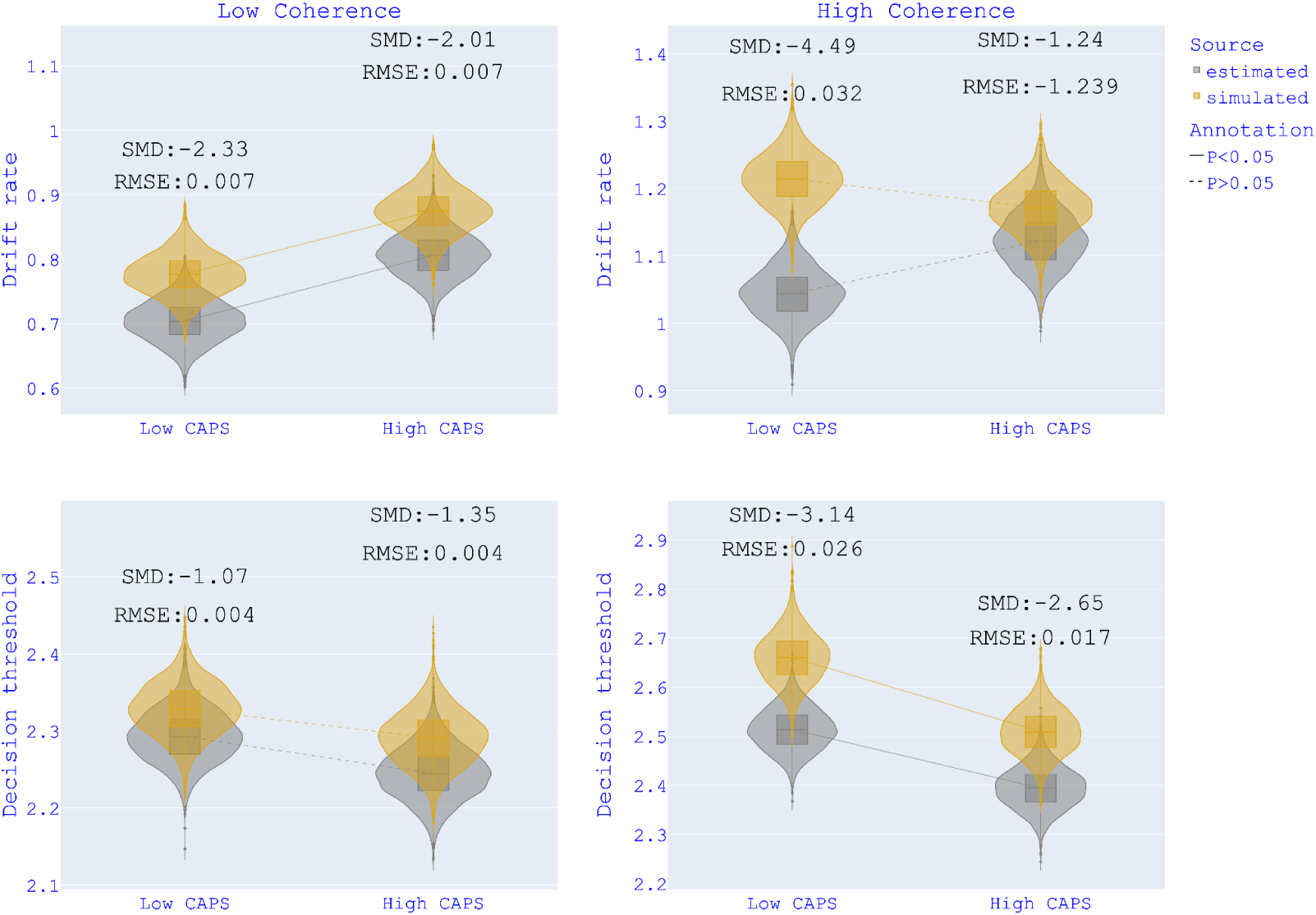
Parameter recovery HDDM-CAPS-C. Recovery of drift rates and decision thresholds for HDDM-CAPS-C (motion-coherence - CAPS groups). The ends of the boxes show the lower and upper quartiles of the distributions, while the line inside the box marks the median. SMD = Standardised mean difference; RMSE = Root mean squared error.

##### Regression HDDMs

In *Figure S5*, we show the quantiles of simulated RT distributions against the observed RT quantiles for the winning regression models.

The simulated models reproduced the positive effect of PDI on drift rate (see *Figure-S6* top-left). While the recovered effects of CAPS only nonsignificantly trended in the same direction as the effects of CAPS shown by the participants, the recovered effects of PDI more fully reproduced the pattern of participants’ behaviour(*Figure S6* right). For the effect of motion-coherence, both the effects on drift rate and decision threshold were well recovered in both models (see *Figure-S7*). The effects of psychotic phenotypes (PDI and CAPS) on DDM parameters were better recovered in the between-group models than in the regressions. However, overall, the RT predictions of regression HDDMs were as good as the ones from between-group HDDMs, which led us to report results from both modelling approaches.

**Fig. S5.**
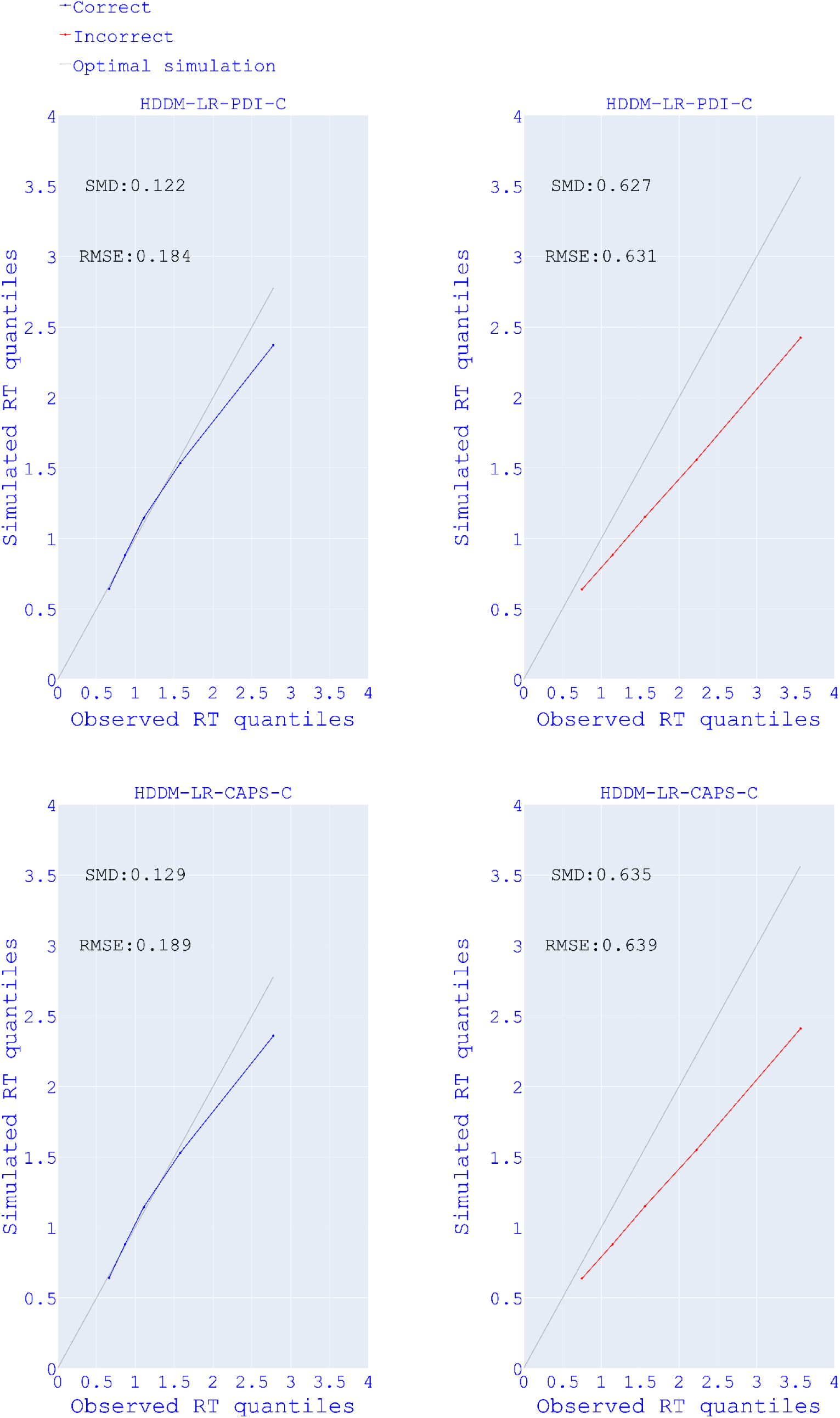
RT predictions of regression HDDMs Study 1. Linearity plots of observed reaction time (RT) quantiles (x-axis) and simulated RT quantiles (y-axis) for correct and error responses of regression models in *Study 1*. SMD = Standardised mean difference; RMSE = Root mean squared error.

**Fig. S6.**
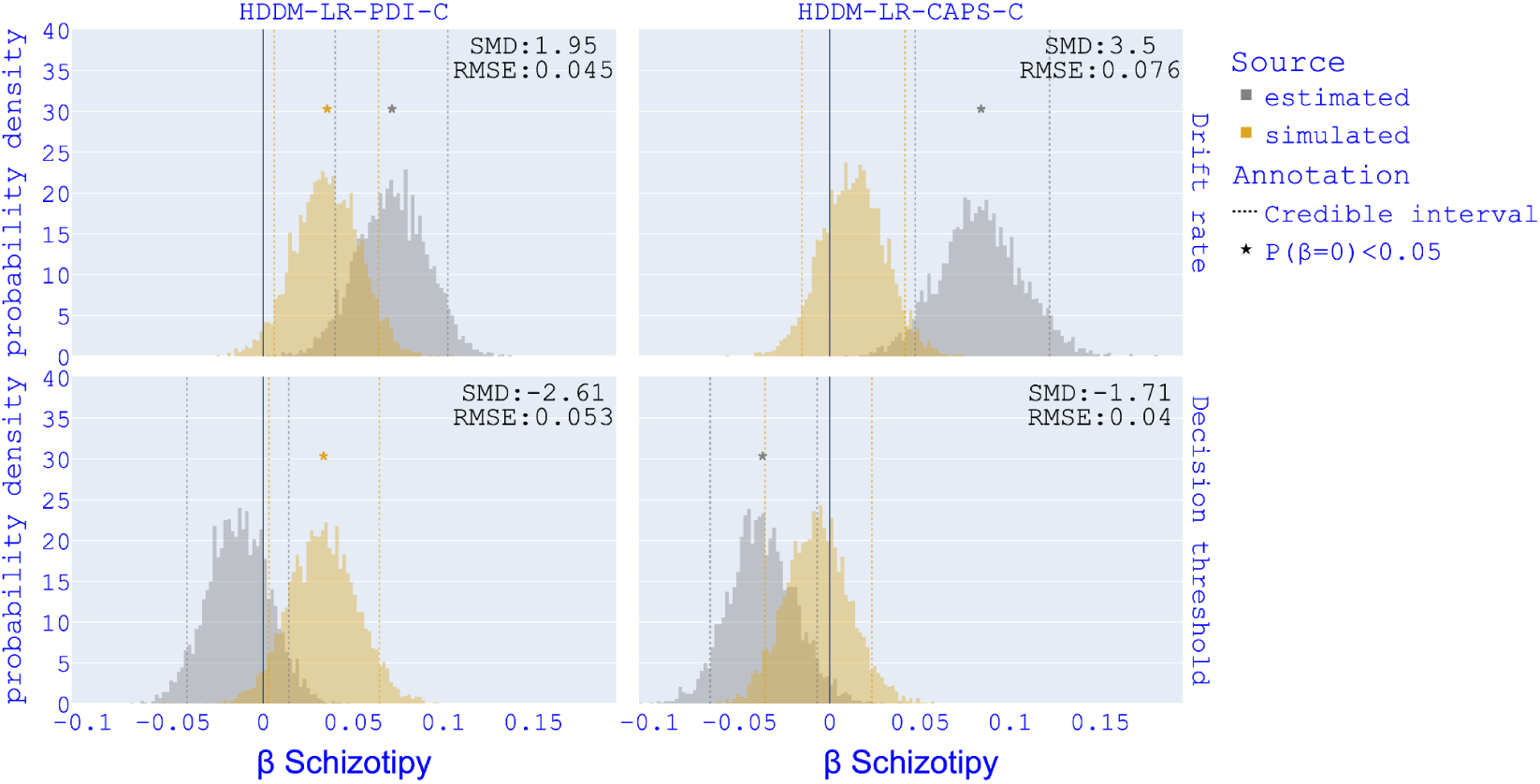
Parameter recovery for psychotic phenotype HDDM-LR-PDI-C, HDDM-LR-CAPS-C. Recovery of β for Schizotopy in HDDM-LR-PDI-C (motion-coherence - PDI groups), HDDM-LR-CAPS-C (motion-coherence - CAPS groups). SMD = Standardised Mean Difference; RMSE = Root mean squared error.

**Fig. S7.**
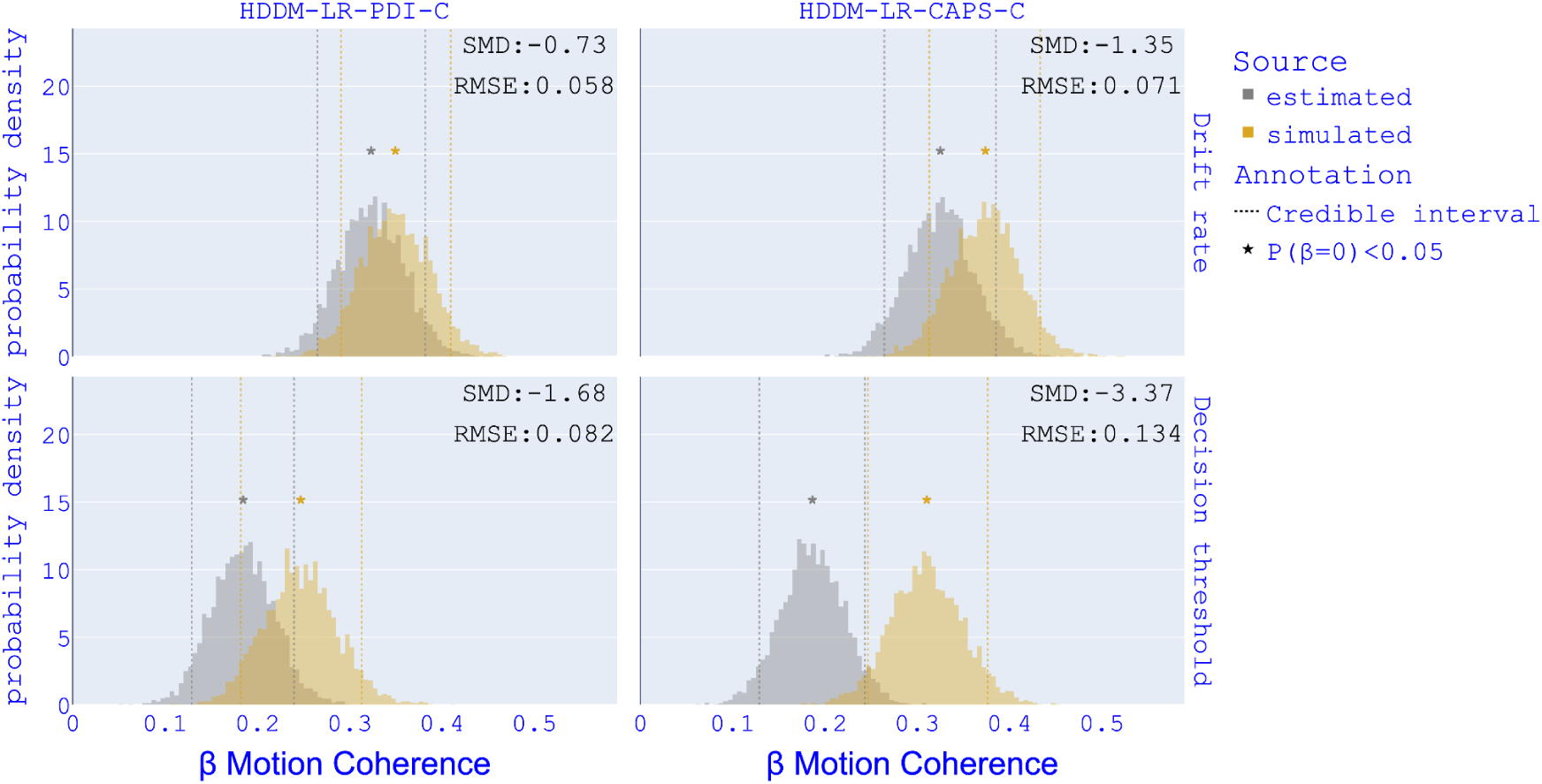
Parameter recovery for motion-coherence HDDM-LR-PDI-C, HDDM-LR-CAPS-C. Recovery of β for motion-coherence in HDDM-LR-PDI-C (motion-coherence - PDI groups), HDDM-LR-CAPS-C (motion-coherence - CAPS groups). SMD = Standardised Mean Difference; RMSE = Root mean squared error.

#### 2.3 Discussion

We performed data simulation and parameter recovery of the HDDMs and evaluated to what extent our models were able to capture the hidden causes that produced our data. We specifically looked at whether our models were able to simulate the empirical data and recover the effects identified in the empirical models within the simulated sample. We recognised five common patterns that we will comment on before going into the specifics of the models.

(1) From the comparison between simulated and empirical RT quantiles, we notice a systematic difference in models’ predictions for correct and error responses. In fact, in simple perceptual decision-making tasks (as the RDM), correct responses outnumber to a great extent error responses, as exemplified by the RT distribution in *Figure*-*2.* With more data available, the models were hence better at predicting RT for correct responses than for error responses. In future experiments, the collection of larger samples might improve predictions of RT for error responses.
(2) Generally, all models underestimated RTs. This prediction error is mainly driven by an underestimation of slow RTs and increases for slower RTs (see *Figure S3* and *Figure S5*).
(3) Between-group models overestimated drift rates. This overestimation seems to be a systematic bias of the models that affected median-split groups uniformly and did not prevent the good recovery of the effects of grouping on the parameter. This overestimation might have produced in the simulated data the aforementioned underestimation of RTs, as RTs are inversely related to the drift rate.
(4) In one case, the simulation models produced what we can call “synthetic false positives” (see *Figure S6* bottom-left), meaning significant effects that were not present in the empirical models. Synthetic false positives could potentially indicate a lack of reliability in the null effects found in the empirical models, but it is uncertain if they can represent actual false negatives in the empirical results.

Although drift rates have been generally overestimated by the models, the simulations reproduced the differences in drift rate between the median-split groups present in the empirical data and reported in the *Results* section. Simulated estimates of decision threshold were also generally higher than the empirical estimates. This overestimation did not prevent the recovery of the negative effect of CAPS on decision threshold.

Regression HDDMs were as good as between-group HDDMs at predicting empirical RTs (see *Figure S3* and *Figure S5*) and recovered all the effects of motion-coherence on DDM parameters. Nevertheless, for psychotic phenotypes, only the negative effect of PDI on the drift rate was well-recovered by regression HDDMs. The trending but nonsignificant CAPS effects in the simulated regression HDDMs presents us with a scenario where we see two types of models, the between-(median-split)-group and regression HDDMs, where the same CAPS effect was detected in the empirical data in both types of HDDM but was in one case well recovered (between-group HDDM) but less so in the other (regression HDDM). This suggests that the associations between DDM parameters and CAPS are present in the data but are better captured by models that make the parameters vary between median-split groups rather than linearly.

### 3. Results of preregistered analysis

*HDDM*. We report here the results from hierarchical drift-diffusion models (HDDMs) we preregistered. The models we preregistered assessed the between-group effects of psychotic phenotypes regardless of motion coherence in all trials (HDDM-1, HDDM-2, HDDM-3), only in trials with 15% motion-coherence (HC-HDDM-1, HC-HDDM-2, HC-HDDM-3) and only in trials with 25% motion-coherence (EC-HDDM-1, EC-HDDM-2, EC-HDDM-3). However, these models could not account for between-participant psychotic phenotype and within-participant variability across different motion-coherence conditions, potentially leading to less precise parameter estimation compared to models we presented in the paper that accounted for both motion-coherence and psychotic phenotype effect at the same time. These preregistered models showed results consistent with the ones presented in the main body of the paper. The model where parameters varied for CAPS group for all trials (HDDM-2) showed a DIC significantly lower relative to the null-model (See *Table S1*). In *Figure S8* we show the results from HDDM-2 that correspond to the results from HDDM-CAPS-C that we showed in the main body of the paper. In the models where only one condition-type trials were used, none of the models showed a DIC value difference higher than 10 compared to the relative null-model (see *Table S2* and *Table S3*), although EC-HDDM-2 (CAPS groups for 25% motion coherence trials) reached a DIC difference of 9.4 (see *Table S3*).

**Table S1.**
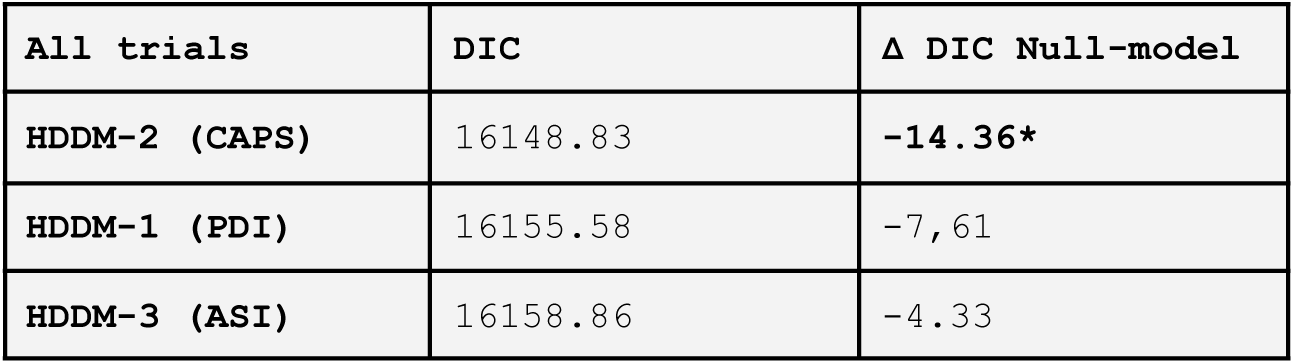
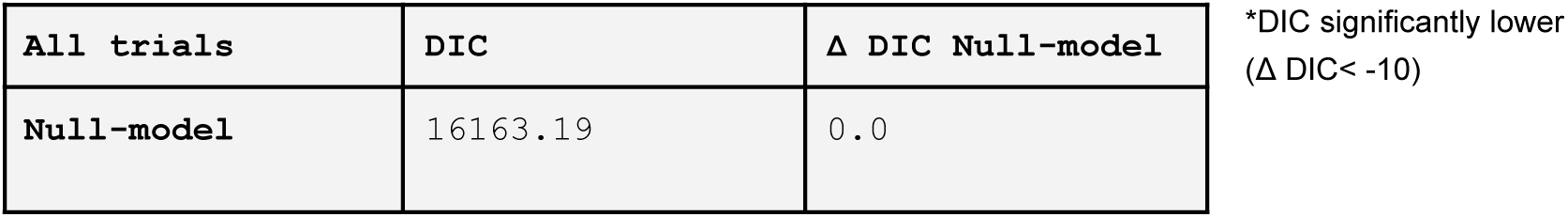
DIC between-group HDDMs. *Values of Deviance Information Criterion (DIC) for each model in descending order (lower DIC, better fit); differences in DIC (*Δ *DIC) between each model and the* Null-model (parameters not varying for any variable).

**Fig S8.**
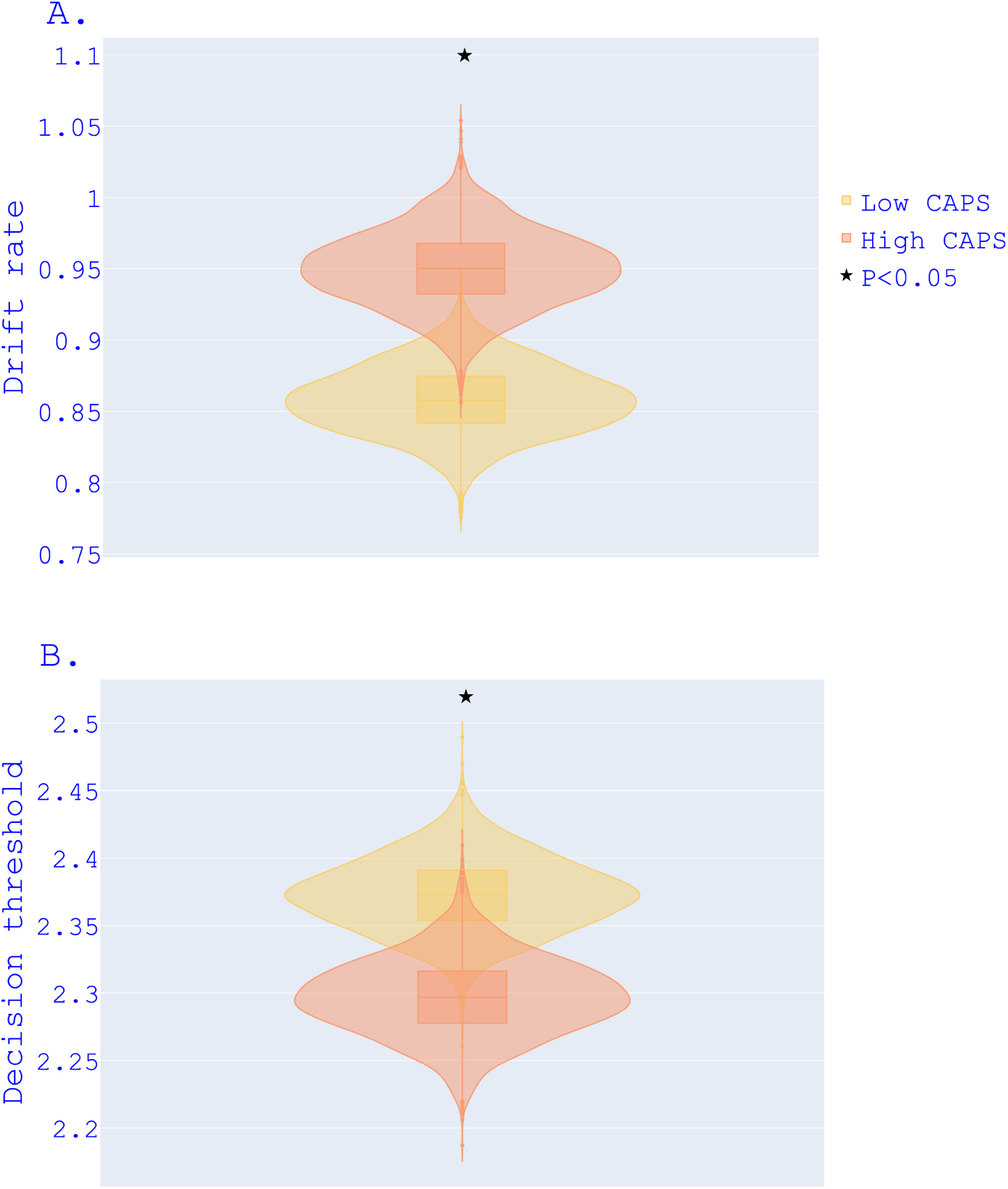
Between-group comparison of winning preregistered model HDDM-2. Posterior probability density distributions of drift rate (A) and decision threshold from HDDM-2 for high-CAPS and low-CAPS groups. The ends of the boxes show the lower and upper quartiles of the distributions, while the line inside the box marks the median.

**Table S2.** DIC between-group HDDMs. *motion coherence=15%. Values of Deviance Information Criterion (DIC) for each model in descending order (lower DIC, better fit); differences in* DIC (Δ *DIC) between each model and* the Null-model (parameters not varying for any variable).

| 15% motion coherence | DIC | $\Delta$ DIC Null-model |
| --- | --- | --- |
| HC-HDDM-2 (CAPS) | 8680.42 | -4.17 |
| HC-HDDM-1 (PDI) | 8681.56 | -3.03 |
| Null-model-15 | 8684.59 | 0.0 |
| HC-HDDM-3 (ASI) | 8686.09 | 1.5 |

\*DIC significantly lower ( $\Delta$ DIC < -10)

**Table S3.** DIC between-group HDDMs. *motion coherence=25%. Values of Deviance Information Criterion (DIC) for each model in descending order (lower DIC, better fit); differences in DIC (*Δ *DIC) between each model and the* Null-model (parameters not varying for any variable).

| 25% motion coherence | DIC | $\Delta$ DIC Null-model |
| --- | --- | --- |
| EC-HDDM-2 (CAPS) | 7388.06 | -9.4 |
| EC-HDDM-3 (ASI) | 7394.42 | -3.04 |
| EC-HDDM-1 (PDI) | 7397.18 | -0.28 |
| Null-model-25 | 7397.46 | 0.0 |

\*DIC significantly lower ( $\Delta$ DIC < -10)

#### Linear regression

Here, we show the results from the ordinary least squares models we preregistered (see *Fig 2*). Because of the violation of the assumption of normal distribution of outcome variables, we did not include them in the main body of the paper. These results do not vary from the quantile regression results reported in the main body of the paper except for the PDI association with Conf-N which here does not reach significance.

**Fig S9.**
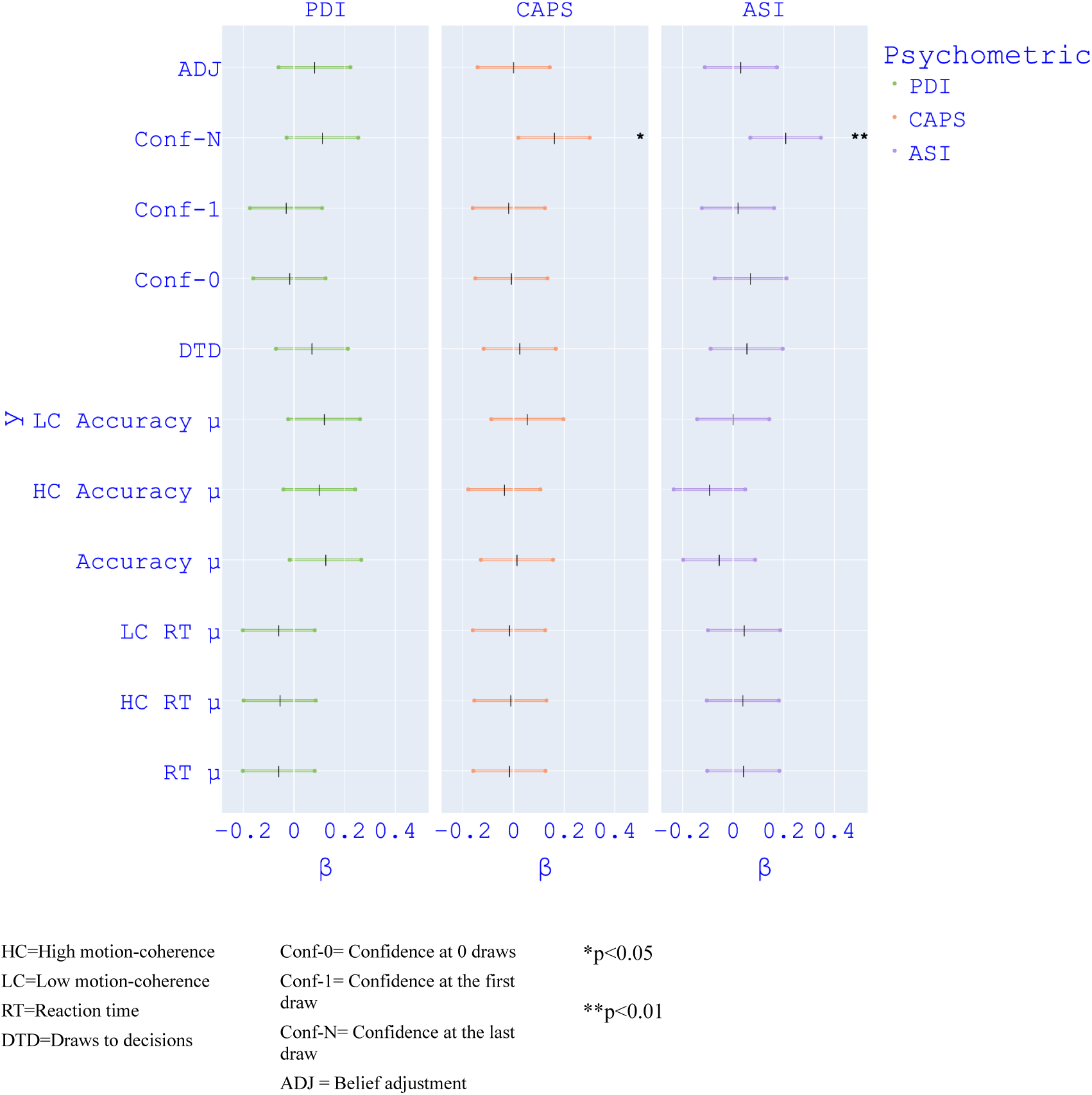
OLS regression models. Results from ordinary least squares models, each with PDI, CAPS and ASI as the predictor. Coloured lines indicate the estimates for *β*.

